# Detection of autoantibodies against the acetylcholine receptor, evaluation of commercially available methodologies: fixed Cell-Based Assay, Radioimmunoprecipitation Assay and Enzyme-Linked Immunosorbent Assay

**DOI:** 10.1101/2023.07.30.23293388

**Authors:** Larissa Diogenes, Alessandra Dellavance, Danielle Cristiane Baldo, Sarah Cristina Gozzi-Silva, Kethellen Gomes, Monica Simon Prado, Luis Eduardo C. Andrade, Gerson Dierley Keppeke

## Abstract

**Introduction:** Myasthenia Gravis (MG) is an autoimmune disease resulting from the action of pathogenic autoantibodies (AAbs) directed against nicotinic acetylcholine receptors (AChR), which interfere with communication between the neurotransmitter acetylcholine and its receptor on the muscle fiber. The detection of anti-AChR using Radio Immuno Precipitation Assay (RIPA) has 100% specificity for the diagnosis of MG, however RIPA has high execution and interpretation complexity and requires radioactive materials, which restrict their use to specialized laboratories.

**Objective:** We compared the performance of the gold standard RIPA with different non-RIPA anti-AChR immunoassays, including a cell-based assay (CBA) and two solid-phase ELISA kits.

**Results:** 145 samples were included with medical indication for anti-AChR testing. By the RIPA method, 63 were negative (RIPA-Neg <0.02 nmol/L), 17 were classified as B*orderline* (≥0.02 – 1 nmol/L), and 65 were positive (RIPA-Pos >1 nmol/L). The competitive ELISA yielded a poor performance with low Kappa agreement with RIPA (0.210). The indirect ELISA yielded a substantial Kappa agreement (Kappa=0.652), with ∼70% sensitivity and ∼96% specificity, compared to RIPA. In a semiquantitative analysis, there was a good Spearman correlation between the indirect ELISA and RIPA levels (r=0.845). The best performance was observed with the CBA that uses fixed cells expressing clustered AChR as antigenic substrate. There was an almost perfect agreement with RIPA (Kappa = 0.969), with ∼97% sensitivity and 100% specificity. However, in the *Borderline* group, only 5 (∼30%) were positive using the CBA method, suggesting a slightly lower sensitivity for the CBA.

**Conclusion:** For detection of anti-AChR reactivity, the indirect immunofluorescence assay yielded a very good analytical performance taking RIPA as the reference method, with potential to replace the RIPA in the clinical laboratory. ELISA could be an option to estimate anti-AChR AAb levels after confirming positivity by the CBA.

## 1. Introduction

Autoantibodies (AAb) are immunoglobulins directed against self-antigens and they are valuable biomarkers for autoimmune diseases, playing a relevant role in the clinical diagnosis and management of several of these diseases. Some AAb are directly involved in the pathophysiology of some autoimmune diseases. Myasthenia Gravis (MG) is a neuromuscular autoimmune disease characterized by muscle weakness and fatigue, resulting from AAb directed mostly (∼85% of cases) against the nicotinic acetylcholine receptor (referred to in this manuscript as AChR or nAChR) present at the muscle cell membrane [1, 2]. Anti-AChR AAb hinder the communication between the nerve and muscle fiber. MG can affect any muscle and can be life threatening when swallowing and breathing are impaired. Although there is no cure for MG, appropriate treatment with acetylcholinesterase-inhibitor as well as immunosuppressive drugs [3-5], or more recently immunobiologicals [6], can somewhat improve the patient’s quality of life and prognosis.

In humans, the muscular nAChR exists as an embryonic and an adult isoform, with the adult form predominating after birth. The embryonic form consists of five subunits, α1, β1, γ, and δ, with a proportion of 2:1:1:1, respectively, arranged in a circular manner to form the cation channel. The adult form is similar but contains the ε subunit instead of the γ subunit [7]. The anti-AChR AAb targets preferentially the extracellular part of the α1 subunit, where ACh binds to promote the “opening” of the cation channel [1, 7].

Circulating anti-AChR AAb has 100% specificity for the diagnosis of MG. The laboratory platforms most commonly applied for the detection of anti-AChR AAb are radioimmunoprecipitation assay (RIPA) and solid-phase immunoassays, such as the enzymatic (ELISA) or chemiluminescent (CLIA) types, for which there are available commercial kits. RIPA was developed in the 1970’s and is considered the gold standard [8, 9] for anti-AChR AAb determination. It is based on a mixture of nAChR from various sources, but mainly from human medulloblastoma TE671 cells [10], with its ligand α-bungarotoxin conjugated to the radioactive isotope ^125^I (iodine-125). After incubation with the patient’s serum containing anti-AChR AAb, the complex is precipitated with a second anti-human IgG antibody and the radioactivity is quantified by a gamma counter [2, 11]. RIPA is highly sensitive but presents the inherent disadvantage of requiring radioactive materials, which makes its execution costly and restrict to a few centers worldwide. Thus, many clinical labs use solid phase immunoassays, such as ELISA and its variations, which have a poor reputation regarding sensibility [12], probably because the native conformation of AChR epitopes are variably degraded during antigen purification and/or coating of the solid-phase plates. This is especially relevant because the AChR is a membrane complex with multiple subunits and anti-AChR AAb bind preferentially to conformational and discontinuous epitopes [13].

Thus, over the past decade if has been proposed the use of cell-based assays (CBA) to detect anti-AChR AAb [14]. CBA is an indirect immunofluorescence assay (IIF) that uses cells transfected or transduced with plasmids that encode the protein of interest. Usually, the transfected gene has a cell membrane localizing sequence that ensures the expression of high levels of the native folded protein at the cell membrane, providing an optimal exposure of the relevant epitopes to be targeted by the autoantibodies. Some studies have reported that anti-AChR immunoassays with live cells expressing clustered nAChR can have a sensitivity equal to or even higher that the RIPA [15-18]. However, the maintenance of live-cell cultures expressing AChR requires special facilities and expertise, which also restricts the adoption of this methodology in most clinical laboratories.

Recently a biochip CBA with four fixed transfected cell-lines has been marketed (Euroimmun). The biochip configuration has cells expressing the adult AChR-ε, the embryonic AChR-γ, muscle-specific kinase (MuSK), and wild-type cells (EU-90) to be used as negative control in the IFA. The two cell-lines expressing the AChR also contain the other subunits as well as rapsyn, a molecule that clusters the receptors, which improves the sensitivity to detect anti-AChR AAb in MG [17, 18]. So far this novel immunoassay has been utilized in only a few studies [14, 19, 20], but it has presented an almost perfect Kappa agreement with the RIPA to detect anti-AChR in MG patients. For simplicity, in this study we will refer to this assay with fixed cells as CBA.

Our goal in this study was to compare the performance of different commercial assays to detect anti-AChR AAbs: the gold standard RIPA, a recently marketed cell-based assay (CBA), and two solid-phase ELISA kits.

## 2. Results

The concentration of anti-AChR Abs measured by the RIPA is given in nmol/L, and various laboratories and assay manufacturers, as well as the Mayo Clinic where our samples were tested, consider results <0.02 nmol/L as non-reactive for anti-AChR, or negative (for details, consult <www.mayocliniclabs.com>) [21]. Thus, among the 145 samples included in the study, 63 were non-reactive for anti-AChR by the RIPA. However, some manufacturers of anti-AChR RIPA kits, such as the RSR (Cardiff, UK), Cisbio Bioassays (France) and DIAsource ImmunoAssays (Belgium), to cite a few examples, recommend a cutoff of >0.4 or 0.5 nmol/L to be consider as positive. Moreover, some publications have recommended a higher cut-off for definition of positive results, as those <1nmol/L may not be true positives [17, 19]. We then classified our results into three groups: 1) <0.02nmol/L as negative (n=63, 43.5%) (RIPA-Neg group); 2) between 0.02 and 1nmol/L as *borderline* (n=17, 11.7%), and 3) >1nmol/L as positives (n=65, 44.8%) (RIPA-Pos group). These three groups were considered the reference for anti-AChR reactivity for evaluation of the performance of the other methods throughout the study (Table 1).

**Table 1.** Reactivity to AChR and HEp-2 cells.

| RIPA<br>(n=145) |  | Negative<br>(n=63)<br><0.02 nmol/L | Borderline<br>(n=17)<br>0.22 (±0.2)<br>nmol/L | Positive<br>(n=65)<br>20.1 (±20.3)<br>nmol/L | p<br>value | Kappa<br>(95% confidence<br>interval)<br>(f) |
| --- | --- | --- | --- | --- | --- | --- |
| Indirect ELISA (n=128) |  | (n=51) | (n=15) | (n=62) |  |  |
| Cutoff (a) | - | 68.6% (n=35) | 53.3% (n=8) | 1.6% (n=1) | <0.001 | 0.688<br>(0.557-0.819) |
|  | + | 31.4% (n=16) | 46.7% (n=7) | 98.4% (n=61) |  |  |
| Cutoff (b) | - | 96.1% (n=49) | 93.3% (n=14) | 29.1% (n=18) | <0.001 | 0.652<br>(0.519-0.785) |
|  | + | 3.9% (n=2) | 6.7% (n=1) | 70.9% (n=44) |  |  |
| Competitive ELISA<br>(c) (n=145) | - | 87.3% (n=55) | 88.2% (n=15) | 66.2% (n=43) | 0.008 | 0.210<br>(0.068-0.351) |
|  | + | 12.7% (n=8) | 11.8% (n=2) | 33.8% (n=22) |  |  |
| CBA<br>(n=145) | AChR-ε | 0% (n=0) | 29.4% (n=5) | 96.9% (n=63) | <0.001 | 0.969<br>(0.928-1.000) |
|  | AChR-γ | 0% (n=0) | 23.5% (n=4) | 90.8% (n=59) |  | 0.909<br>(0.839-0.978) |
|  | CBA-ε/γ-Neg | 100% (n=63) | 70.6% (n=12) | 3.1% (n=2) |  | - |
| HEp-2 IFA | Neg, 48.9%<br>(n=71) | 47.6% (n=30) | 41.2% (n=7) | 52.3% (n=34) | 0.687 | - |
|  | Pos, 51.1%<br>(n=74) | 52.4% (n=33) | 58.8% (n=10) | 47.7% (n=31) |  |  |
| Titer (d) |  | 1/467 (±483) | 1/271 (±162) | 1/415 (±339) | 0.489# | - |
| Patterns<br>(ICAP)<br>(e) | Nuclear speckled<br>(AC-2, AC-4, AC-5) | 23 (59%) | 8 (62%) | 22 (63%) | 0.948 | - |
|  | Nucleolar<br>(AC-8 to AC-10) | 5 (13%) | 1 (8%) | 6 (17%) |  |  |
|  | Cytoplasmic<br>(AC-15 to AC-23) | 7 (18%) | 2 (15%) | 4 (11%) |  |  |
|  | Others (AC-3, AC-7,<br>and AC-25 to AC-28) | 4 (10%) | 2 (15%) | 3 (9%) |  |  |

➤ data is presented as % for categorical variables, or averages ± S.D in quantitative variables (d).
(a) Cutoff Youden index J = 0.8409;
(b) Cutoff calculated as the average + 3 S.D. of the RIPA-Neg group = 4.568;
(c) Cutoff calculated as the average + 1 S.D. of the RIPA-Neg group = 20 ng/mL;
(d) HEp-2 IFA titer was compared among the positive samples for this test (# Kruskal-Wallis test);
(e) Some samples presented more than one pattern (ICAP nomenclature);
(f) To quantify the agreement with Cohen's Kappa, only the RIPA negative/positive groups were compared with the negative/positive in the ELISAs and in the CBA, respectively.

Anti-AChR reactivity was evaluated in 128 samples by indirect ELISA and in 145 samples by competitive ELISA. Among the 128 samples tested in the indirect ELISA, 62 were positive (RIPA-pos) and 51 were negative (RIPA-neg) in the RIPA assay. Using the cutoff recommended by the manufacturer of the indirect ELISA (≥0.5 nmol/L as positives), there was a high proportion of false positives in the RIPA-Neg group (43%, 21 out of the 51 samples), thus we first adjusted the cutoff based in the Youden index J (0.84 nmol/L) [22]. This method for calculating cutoff usually promotes the sensitivity in a given assay [23] (Figure 1A). From the 62 RIPA-Pos samples, 61 (98.4%) were considered positive by the indirect ELISA with the Youden J cutoff, whereas from the 51 RIPA-Neg samples, only 35 were also considered negative in the indirect ELISA, yielding a specificity of 68.6% (95% CI 54.9%-79.6%) (Table 2). There was a substantial agreement between RIPA and the indirect ELISA with the adjusted cutoff (Kappa = 0.688, 95% CI 0.557-0.819) (Table 1).

**Table 2.** Sensitivity and Specificity for the anti-AChR immunoassays in comparison with RIPA.

|  | RIPA-positive and RIPA-Negative samples (e) |  |  |
| --- | --- | --- | --- |
|  |  | Value | 95% CI |
| Indirect ELISA (a) | Sensitivity | 0.983 | 0.914-0.999 |
|  | Specificity | 0.686 | 0.549-0.796 |
|  | PPV | 0.792 | 0.688-0.867 |
|  | NPV | 0.972 | 0.858-0.998 |
| Indirect ELISA (b) | Sensitivity | 0.709 | 0.587-0.807 |
|  | Specificity | 0.960 | 0.867-0.993 |
|  | PPV | 0.956 | 0.854-0.992 |
|  | NPV | 0.731 | 0.614-0.822 |
| Competitive ELISA (c) | Sensitivity | 0.338 | 0.235-0.459 |
|  | Specificity | 0.873 | 0.768-0.934 |
|  | PPV | 0.733 | 0.555-0.858 |
|  | NPV | 0.561 | 0.462-0.655 |
| CBA - AChR-ε (d) | Sensitivity | 0.969 | 0.894-0.994 |
|  | Specificity | 1.000 | 0.945-1.000 |
|  | PPV | 1.000 | 0.945-1.000 |
|  | NPV | 0.969 | 0.894-0.994 |

**PPV** = Positive Predictive Value
**NPV** = Negative Predictive Value
(a) Cutoff Youden index J = 0.8409;
(b) Cutoff Average of the RIPA-Neg group + 3 S.D. = 4.568;
(c) Cutoff Average of the RIPA-Neg group + 1 S.D. = 20 ng/mL;
(d) Only results for CBA-AChR- $\epsilon$ were considered for this analysis, and not the CBA-AChR- $\gamma$ ;
(e) Only the groups RIPA positive and Negative were considered, and not the *Borderline*.

**Figure 1.**
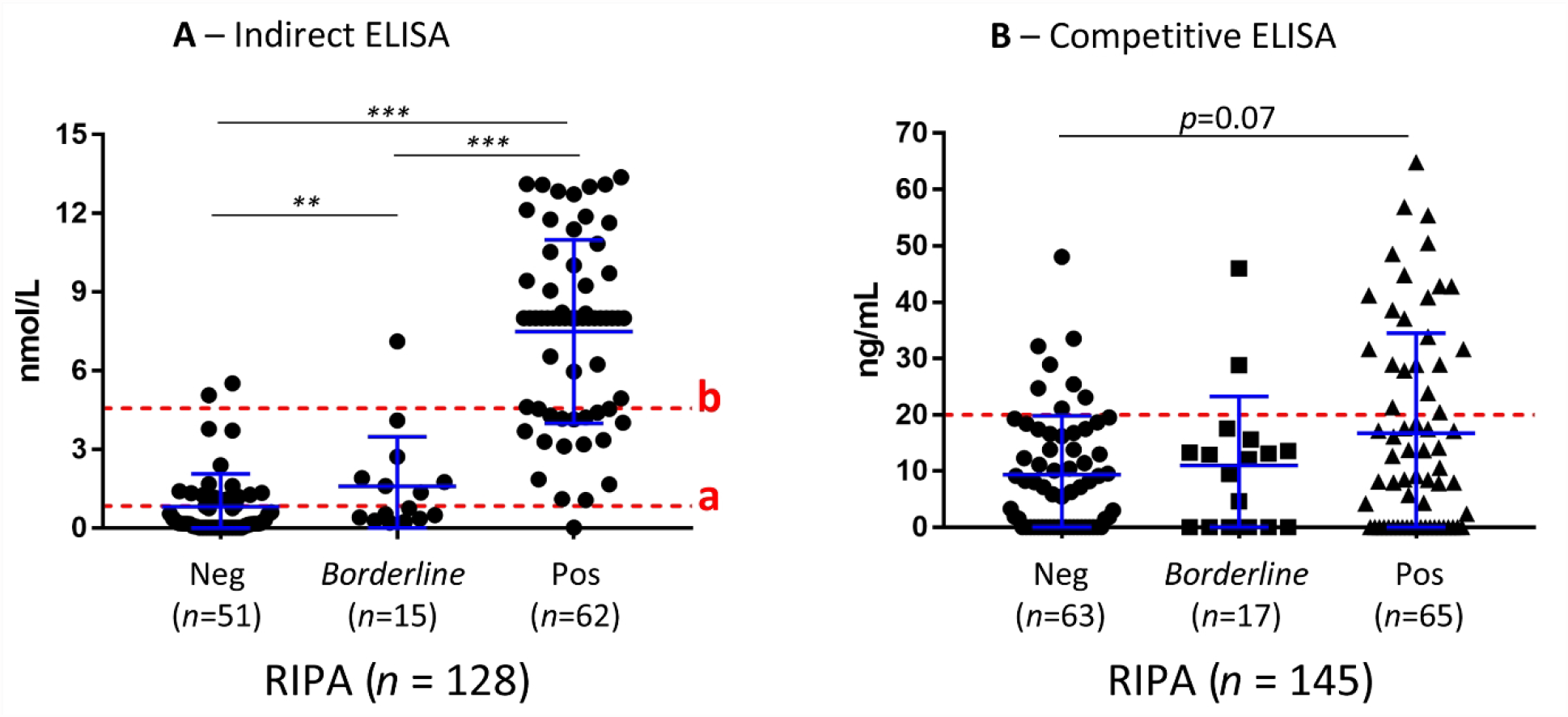
Anti-nAChR reactivity by ELISA. (A) Anti-nAChR reactivity analyzed using an indirect ELISA. Line “a” indicates the Youden index J cutoff = 0.8409, that promotes sensitivity. Line “b” indicates a cutoff that promotes specificity = 4.568, calculated based on the average + 3SD of the RIPA-negative group **p≤0.01; ***p≤0.001. Error bars = SD (B) Analysis using a Competitive ELISA. The cutoff of 20ng/mL was based on the average + 1SD of the RIPA-negative group.

Because anti-AChR AAb is considered to be specific for the diagnosis of MG, assay specificity is more important than sensibility [2]. The specificity obtained with the cutoff adjusted using Youden J index was not satisfactory, thus we defined another cutoff (4.56 nmol/L) based on the average + 3*SD (+3 times the Standard Deviation) of the RIPA-Neg samples (Figure 1A), this method for stablishing the cutoff promotes the specificity in a given assay. With this cutoff, 44 of the 62 RIPA-Pos samples (70.9%) were considered positive and 49 (96.1%) of the 51 RIPA-Neg samples were considered negative in the indirect ELISA, yielding a specificity of 96.0% (95% CI 86.7%-99.3%) (Table 2) and substantial agreement with RIPA (Kappa = 0.652, 95% CI 0.519-0.785) (Table 1).

The competitive ELISA gives results for anti-AChR reactivity in ng/mL and the sensitivity of the assay is ≥1 ng/mL. Curiously, there was not a statistical difference in reactivity between the RIPA-pos and RIPA-neg groups, although the p value (p=0.07) indicated a trend for higher anti-AChR AAb concentration in the RIPA-Pos (Figure 1B). The manufacturer does not suggest a positive/negative cutoff, thus for this study we defined the cutoff based in the RIPA-Neg average +1*SD (20 ng/mL). The proportion of positives by the competitive ELISA was higher in the RIPA-Pos group when compared to the RIPA-Neg (33.8% versus 12.7%, respectively, p=0.008), but there was only a fair agreement rate between the competitive ELISA and the RIPA (Kappa = 0.210, 95% CI 0.068-0.351) (Table 1). With this assay, although specificity was satisfactory (87.3%, 95% CI 76.8%-93.4%), sensitivity was poor (33.8%, 95% CI 23.5%-45.9%) (Table 2).

The samples were then tested with the fixed CBA biochip, which can individually detect reactivity to the adult AChR-ε as well as the embryonic AChR-γ, visualized by the staining signal in the membrane of the cells expressing AChR (Figure 2). Four samples showed reactivity only against the adult AChR-ε isoform (example in Figure 2C), and the additional 59 CBA-positive samples showed reactivity against both isoforms (Figure 2A and B) meaning none of the samples showed reactivity only against the AChR-γ isoform (Table 1). Among the 65 RIPA-Pos samples, 63 (96.9%) were positive in the fixed CBA, yielding a sensitivity of 96.9% (95% CI 89.4%-99.4%) (Table 2). In addition, none of the 63 RIPA-Neg samples showed reactivity in the fixed CBA, meaning 100% specificity for this assay (100.00%, 95% CI 94.5%-100.0%) (Table 2). There was an almost perfect agreement between the RIPA and the fixed CBA (Kappa=0.969, 95% CI 0.928-1.000). Among the 17 RIPA-*Borderline* samples five (29.4%) were positive in the fixed CBA (Table 1).

**Figure 2.**
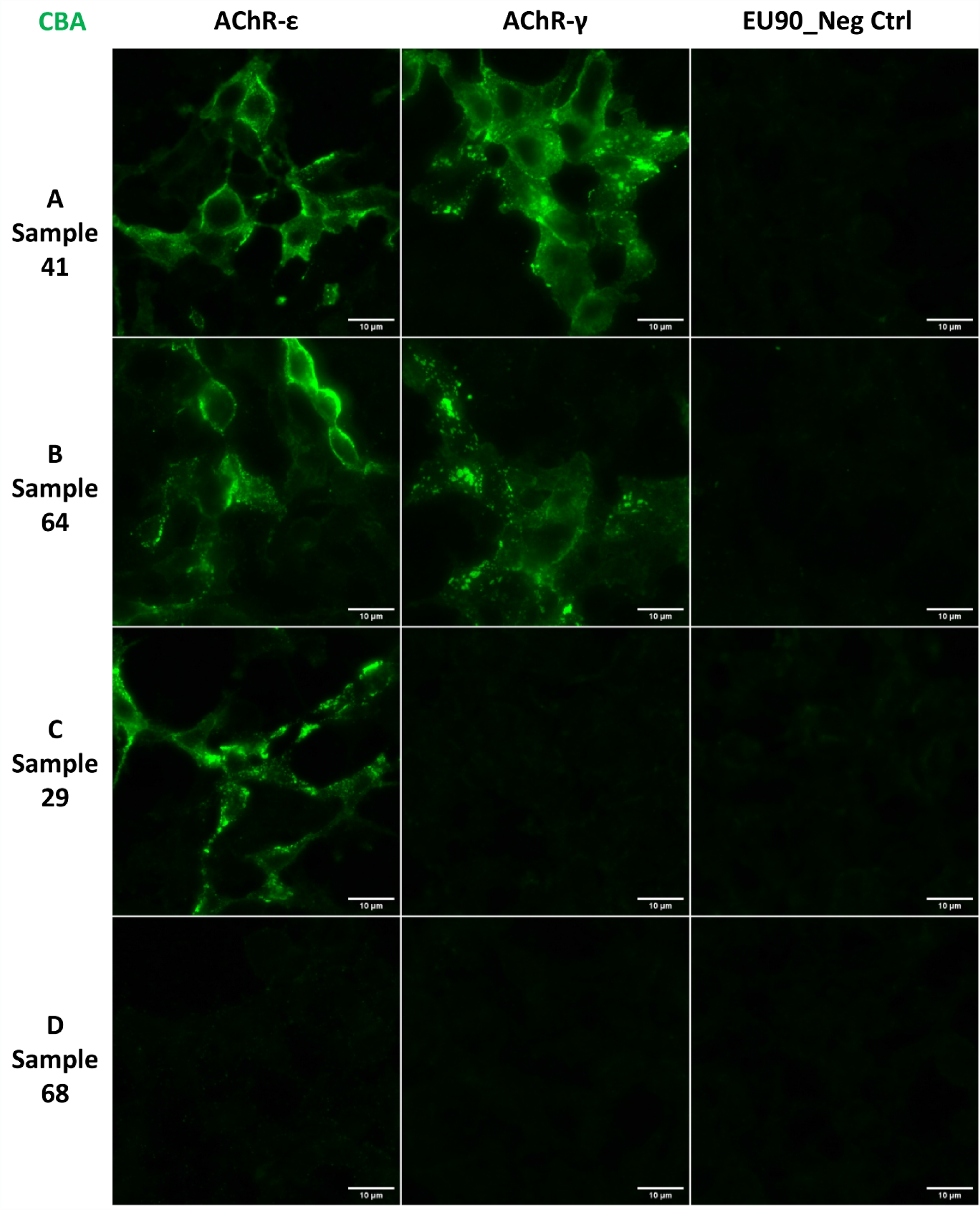
Anti-nAChR reactivity by the fixed Cell-Based Assay (CBA). Indirect Immunofluorescence analysis with EU90 cells expressing AChR-ε or AChR-γ. (A and B) Examples of samples with reactivity for ε and γ. (C) Example of a sample with reactivity only for ε. (D) Example of a sample without any reactivity. Scale bars = 10μm.

Because the fixed CBA is an IIF assay, we investigated how the presence of other AAbs would affect the anti-AChR reactivity and interpretation/visualization of the fixed cells at the microscope. Thus, all samples were tested for anti-cell antibody (antinuclear antibody) by the HEp-2 indirect immunofluorescence assay (HEp-2 IFA). The proportion of anti-cell reactivity was similar among the RIPA-Neg, RIPA-Pos and *Borderline* samples (p=0.687), with about ∼50% positivity in all RIPA groups (Table 1), suggesting that other AAbs in given samples do not interfere with interpretation of anti-AChR reactivity in the CBA. Of special interest, samples with HEp-2 IFA with cytoplasmic staining could be correctly interpreted regarding reactivity to AChR in the fixed CBA. The distribution of HEp-2 IFA patterns, as well as titers, were also similar among the groups (Table 1).

## 3. Discussion

Anti-AChR antibodies represent the main laboratory parameter for the diagnosis of MG. Due to the practical difficulty in running the gold standard RIPA for anti-AChR in clinical laboratories, ELISA kits have been developed as an alternative approach by the in vitro diagnostic industry. However, detection of anti-AChR autoantibodies for MG diagnosis using ELISA in clinical laboratories has been questioned due to the low accuracy of this technique for this particular autoantibody system [12, 14, 24, 25], and this was not different in our study. Among the two ELISA kits tested, better results were observed with the indirect ELISA with adjusted cutoff that promotes higher specificity (in this case, 96.0% specificity), but at the expense of low sensitivity (∼70% of the RIPA-pos samples). For the competitive ELISA, specificity was satisfactory and sensitivity was poor, making this specific kit impractical for clinical application. Since we only had access to these two ELISA kits, we cannot generalize our findings to other commercially available products that could provide better performance [2]. Employment of signal enhancing strategies, such as those based in avidin/biotin, could also be of benefit, as discussed elsewhere [2].

The recently developed anti-AChR CBA with fixed cells appears as a promising alternative to RIPA to be used in the clinical laboratory. Confirming recent studies in Italian and Canadian cohorts [14, 19, 20], our findings with a Brazilian cohort also showed a good performance of the fixed CBA for detection of anti-AChR when compared to RIPA, with an almost perfect agreement (Kappa=0.969). This assay has the potential to replace RIPA effectively in the clinical laboratory in the detection of anti-AChR AAb. However, we must highlight that the CBA with fixed cells was less sensitive to detect low titer anti-AChR, as only ∼30% of the samples in the RIPA *Borderline* group were positive in the CBA. Although details regarding commercial slides manufacturing are proprietary, the cells are usually dehydrated with alcohol fixatives, to facilitate storage and distribution. Alcohol fixatives precipitate proteins and some of the binding between membrane proteins is lost, as recently demonstrated elsewhere [26]. In the anti-AChR CBA, cell fixing could affect, up to some degree, the receptor integrity and clustering, which would in turn affect autoantibody binding and impair detection of low titer anti-AChR. It has been suggested that live cells expressing clustered nAChR to detect the AAbs can show a sensitivity even higher than RIPA itself, because transfected cells better resembles the physiological expression of the AChR by myocytes [15-18].

Replacement of RIPA has the benefit of avoiding radioactive components and considerably lowering the assay costs. A practical strategy would be the screening of samples in a fixed CBA. Since anti-AChR titers correlate with the clinical course of MG [27, 28], positive samples could be processed for the determination of anti-AChR serum levels by conventional RIPA, ELISA or by titration in fixed CBA. This strategy may first need to be validated by future studies since CBAs can make the use of signal enhancing molecules such as avidin/biotin. In this sense, there was a good correlation between anti-AChR levels by RIPA (65 RIPA-Pos + 17 *Bordeline* samples) and the indirect ELISA results (Spearman r=0.845 (95%CI 0.756-0.903; p=<0.001).

However, as already mentioned, the CBA with fixed cells was less sensitive in samples with low Ab titers. Thus, in case CBA and ELISA yield negative reactivity, but there is strong clinical indication of MG, such as suggestive electroneuromyography results, among others [3], the sample should be further tested using either RIPA or CBA with live cells expressing clustered AChR, demonstrated to have improved sensibility to detect anti-AChR AAbs even at low levels [15, 18, 20].

CBA also has some drawbacks beyond low sensitivity in samples with low AAb titers. A positive reaction on the CBA with fixed cells is given by observing the labeling of the AChR subunits on the membrane of the cells under a microscope. Samples that do not react with the ε or γ subunits are considered negative (Figure 2) [14]. As observed in other studies [19], we noticed that some samples yielded an excessive non-specific background staining, and curiously a portion of those samples were also positive for anti-cell antibody in the HEp-2 IFA (data not shown). Thus, it is important for any CBA to be performed and analyzed at the microscope by trained personnel to avoid erroneous interpretation.

Although the samples included in this study arrived with the medical request (check Materials and methods) to test for the presence of anti-AChR/anti-Musk, meaning it’s likely the patients presented neuromuscular symptoms that supported the request, the major limitation in our study was the impossibility of accessing the patient’s diagnostics and clinical features. This limitation prevented us from knowing if, for example, some of the RIPA-Neg patients would be diagnosed as serum-negative MG (SNMG). We also could not investigate a possible association of reactivity to specific adult or embryonic AChR assemblies with the Ocular and Generalized MG forms. Future studies should address such points.

In summary, the fixed CBA test presented better performance than the two ELISA products and showed an almost perfect agreement (Kappa=0.969) and 100% specificity compared to the gold standard RIPA test. This kit was recently launched commercially and is currently in exclusive use for research purposes, but it has promising potential as an alternative to RIPA in the clinical laboratory, especially due to its radiation-free nature.

## 4. Materials and methods

### 4.1 Samples

A total of 145 samples with medical request for anti-AChR/anti-Musk AAb testing were retrieved from the immunology division at Fleury Group Laboratory, Sao Paulo, Brazil. Because our goal was to study anti-AChR reactivity, samples with anti-Musk reactivity were not included in the study, therefore it was expected that none of the 145 samples would show reactivity to MuSK.

Because the patient’s clinical information was not accessed and the assays performed were as those requested by the patient’s physician, the ethics committee waived the need to collect the patient’s informed consent. The study was approved by the Ethics committee at Fleury Group (Plataforma Brasil CAAE: 57480622.3.0000.5474).

### 4.2 Assays

The following assays were performed in all samples: 1) Anti-AChR by RIPA (Mayo Clinic, USA) on a clinical-service basis; 2) Indirect ELISA (EA 1435-9601 G, Euroimmun, Germany), this kit is registered at the Brazilian regulatory agency to detect anti-AChR (ANVISA 81148560050); 3) Competitive ELISA - Research Use Only (MBS729942, MyBioSource, San Diego, CA, USA); 4) Fixed CBA Myasthenia gravis Mosaic 2 (FA 1435-1010-2, Euroimmun, Germany); 5) Anti-cell antibody (antinuclear antibody) by HEp-2 IFA (FA 1520-2010, Euroimmun, Germany). All assays were performed following the respective manufacturer’s protocol.

Immunofluorescence slides (the CBA and the HEp-2 IFA) were analyzed for positivity and staining patterns in a fluorescence microscope with 200x or 400x magnification (Axio Imager.M2, Carl Zeiss, Germany). Anti-cell antibody titer was determined with sequential double dilutions starting at 1/80 up to end titer.

### 4.3 Data analysis

Quantitative and semi-quantitative parameters were evaluated for normal distribution with D’Agostino & Pearson normality test, and the distribution was not normal for all of them. Thus, when averages of two groups were compared, Mann-Whitney test was applied, when three or more groups were compared, Kruskal-Wallis test was applied. The correlation between anti-AChR levels by ELISA or RIPA was calculated using Spearman test.

Youden J Index for cutoff determination was calculated with MedCalc Statistical Software version 20.115 (MedCalc Software Ltd, Belgium). The proportion of qualitative variables were compared with two-tailed Chi-squared test (Table 1). To calculate sensitivity and specificity of the assays, positive/negative groups were compared in 2x2 tables by Chi-square with Yates’ correction (Table 2).

Cohen’s Kappa agreement was quantified using GraphPad online quickcalcs tool, which uses Fleiss equations to compute the standard error (SE) and confidence intervals. All other analysis were performed using GraphPad Prism v7 (Dotmatics, Boston, MA, USA). *p* values were considered significant when below 0.05, and for all analysis the 95% confidence interval (CI) is also presented when appropriate.

## Data Availability

All data produced in the present study are available upon reasonable request to the authors.

## 5. Author declarations

### Ethical approval

The study was approved by the local Ethics committee at Fleury group (Plataforma Brasil CAAE: 57480622.3.0000.5474).

